# Immunogenicity and Safety of Beta Adjuvanted Recombinant Booster Vaccine

**DOI:** 10.1101/2022.05.25.22274904

**Authors:** Odile Launay, Marine Cachanado, Liem B Luong Nguyen, Laetitia Ninove, Marie Lachâtre, Inès Ben Ghezala, Marc Bardou, Catherine Schmidt-Mutter, Renaud Felten, Karine Lacombe, Laure Surgers, Fabrice Laine, Jean-Sébastien Allain, Elisabeth Botelho-Nevers, Marie-Pierre Tavolacci, Christian Chidiac, Patricia Pavese, Bertrand Dussol, Stéphane Priet, Dominique Deplanque, Amel Touati, Laureen Curci, Eleine Konate, Nadine Ben Hamouda, Anissa Besbes, Eunice Nubret, Florence Capelle, Laurence Berard, Alexandra Rousseau, Eric Tartour, Tabassome Simon, Xavier de Lamballerie, the ARNCOMBI Study Group

## Abstract

**Background:** Variant-adaptated vaccines against coronavirus disease 2019 (COVID-19) as boosters are needed to increase a broader protection against SARS CoV-2 variants. New adjuvanted recombinant protein vaccines as heterologous boosters could maximize the response.

**Methods:** In this randomized, single-blinded, multicenter trial, adults who had received two doses of Pfizer-BioNTech mRNA vaccine (BNT162b2) 3 to7 months before were randomly assigned to receive a boost of BNT162b2, Sanofi/GSK SARS-CoV-2 adjuvanted recombinant protein MV D614 (monovalent parental formulation) or SARS-CoV-2 adjuvanted recombinant protein MV B.1.351 vaccine (monovalent Beta formulation). The primary endpoint was the percentage of subjects with a ≥10-fold increase in neutralizing antibody titers for the Wuhan (D614) and B.1.351 (Beta) SARS-CoV-2 viral strains between day 0 and day 15.

**Findings:** The percentages of participants whose neutralizing antibody titers against the Wuhan (D614) SARS-CoV-2 strain increased by a factor ≥10 between day 0 and day 15 was 55.3% (95% CI 43.4-66.7) in MV D614 group (n=76), 76.1% (64.5-85.4) in MV B.1.351 (Beta) group (n=71) and 63.2% (51.3-73.9) in BNT162b2 group (n=76). These percentages were 44.7% (33.3-56.6), 84.5% (74.0-92.0) and 51.3% (39.6-63.0) for the B.1.351 (Beta) viral strain, respectively. Higher neutralizing antibodies rates against Delta and Omicron BA.1 variants were also elicited after Sanofi/GSK MV Beta vaccine compared to the other vaccines. Comparable reactogenicity profile was observed with the three vaccines.

**Interpretation:** Heterologous boosting with the Sanofi/GSK Beta formulation vaccine resulted in a higher neutralizing antibody response against Beta variant but also the original strain and Delta and Omicron BA.1 variants, compared with mRNA BNT162b2 vaccine or the Sanofi/GSK MVD614 formulation. New vaccines containing Beta spike protein may represent an interesting strategy for broader protection against SARS CoV-2 variants.

**Funding:** French Ministries of Solidarity and Health and Research and Sanofi

**Trial registration number:** ClinicalTrials.gov identifier NCT05124171; EudraCT identifier 2021-004550-33.

## INTRODUCTION

The efficacy of COVID-19 vaccines for reducing the risk of severe COVID-19 infection has now been demonstrated in clinical setting, but also in real-world setting.^1^ However, questions remain regarding the persistence of immunity and potentially less protection against variant viruses.^2^ Therefore, the need for additional doses (boosters) at some interval after the primary vaccination was raised when new waves of COVID-19 cases appeared due to the highly transmissible Delta and Omicron variants.^3^

The matching of vaccines from different platforms for priming and boost was recommended after reports of unusual thrombotic events with the adenovirus-based vector vaccine ChAdOx1-nCov-19 vaccine from AstraZeneca/Oxford.^4^ Several studies have investigated heterologous prime-boost vaccination combining ChAdOx1-nCov-19 for priming and mRNA vaccine (BNT162b2 or mRNA-1273) for boost.^5-7^ In the observational study of Schmidt *et al*, the heterologous vaccine regimen induced higher levels of spike-specific IgG, neutralizing antibodies and spike-specific CD4^+^ T-cells compared to homologous vector vaccine boost and higher or comparable in magnitude to homologous mRNA prime-boost vaccine regimens.^6^

Several vaccines developed more recently might offer an interesting alternative for boost in terms of accessibility, cost, reactogenicity, thermostability and acceptability and could be more immunogenic. The vaccines developed by Sanofi are based on recombinant SARS-CoV-2 spike proteins adjuvanted with GSK AS03.^8^ Two monovalent formulations of this vaccine are currently under development, the first targeting the S protein of the Wuhan (D614) strain and the second targeting the B.1.351 variant (Beta).^9,10^ These vaccines could be of interest as heterologous booster to increase both the magnitude and duration of immune response against the new variants SARS CoV-2. They may also increase the breadth of protection against other variants, such as Omicron, and the formulation including the B.1.351 variant (Beta) spike protein is of specific interest since it has the potential to provide a different spectrum of cross-protection.

The objective of the study was to evaluate the immunogenicity and safety of the adjuvanted recombinant protein vaccine Sanofi/GSK-D614 or -B.1.351 administered as a heterologous booster dose compared to an mRNA vaccine (BNT162b2) administered as a homologous booster dose.

## METHODS

### Study design

We conducted a randomized, single-blinded, multicenter trial across 11 centers in France. Participants were recruited from December 8, 2021, to January 14, 2022.

The objective of the study was to evaluate the immunogenicity and safety of a homologous booster dose of a COVID-19 mRNA vaccine (BNT162b2) or adjuvanted recombinant vaccine with adjuvant (Sanofi/GSK MV(D614) formulation or MV B.1.351 (Beta) formulation) in recipients primed with two doses of BNT162b2 between 3 and 7 months earlier.

The protocol was conducted in accordance with the Declaration of Helsinki and French law for biomedical research. It was approved by the “CPP Ile de France III” Ethics Committee and the French Health Products Safety Agency (ANSM).

This study is registered with the ClinicalTrials.gov identifier NCT05124171 and with the EudraCT identifier 2021-004550-33.

### Participants

Adults aged 18 years and older in good health or with stable health if there was a pre-existing medical history were eligible to participate if they previously received two doses of BNT162b2 with an interval of 3 to 6 weeks and the second dose administered between 3 and 7 months prior to the administration of the study booster dose. Main exclusion criteria were pregnancy or breastfeeding, acute febrile infection within the previous 72 h and/or presenting symptoms suggestive of COVID-19 within the previous 28 days or having been in contact with an infected individual for the last 14 days before the inclusion visit, virologically documented history of COVID-19 (PCR or serology), use of immunosuppressive medications or any immunosuppressive condition that may reduce the immune response, history of severe post-vaccination allergic manifestations or a history of allergic reaction at the time of the first vaccine injection, having received BCG (tuberculosis) vaccine within the previous year or another vaccine within two weeks prior to the boost injection or scheduled to receive a licensed vaccine within 2 weeks after the boost injection.

### Interventions

Participants were randomly assigned in a 1:1:1 ratio to receive BNT162b2, MV(D614) vaccine or MV(Beta) vaccine as a third dose. The Sanofi/GSK adjuvanted recombinant protein vaccines are based on pre-fusion S antigens with the transmembrane domain replaced with a trimerization domain with AS03, an oil-in-water emulsion that contains squalene and α- tocopherol based immunologic adjuvant manufactured by GSK.^11^

Randomization was stratified on center and age group (18–64 years or ≥ 65 years). A web-based randomization system was used (CleanWeb e-CRF, Telemedecine Technologies, S.A.S), with a centralized block randomization list with blocks of size 4 (not communicated to the investigating team). The randomization list was generated by an independent statistician from the trial clinical research unit (URC-EST). Participants were randomized by the investigator.

Vaccines were administered intramuscularly into the deltoid muscle on day 0 by trained personnel. The health care professional administering the vaccine was aware of the treatment group because of differences in the preparation of the vaccines. The injection was therefore performed by a person not involved in the study and the investigating physician did not know which vaccine the volunteer had received. The central laboratories performing the antibody analyses were also blinded to limit measurement bias. Blood samples were planned at D0, D3, D15, D28, D90 and D365 for serological analysis.

### Laboratory assay

Neutralizing antibodies against the Wuhan (D614) SARS-CoV-2 viral strains and B.1.351 (Beta), Delta and Omicron BA.1 variants were assessed with a microneutralization test as previously described.^12^ The test uses clinical strains of SARS-CoV-2 (100 TCID50/well), TMPRSS2-expressing VeroE6 cells and relies on cytopathic effect (CPE) identification at 5 days post-infection. It is a VNT100 (100% of wells lysed in duplicate format). The test is automated in a NSB3 laboratory for all dilution and dispensing steps and for CPE reading. Dilutions tested were 20, 40, 80, 160, 320, 640 and 1280. The range was extended if a titer of 1280 was observed in the first instance.

Anti-SARS-CoV-2 IgG antibodies directed against the S1 domain of the virus Spike protein and the nucleocapsid were assessed using the QuantiVac ELISA kit from Euroimmun^®^ (Lubeck, Germany).

Spike specific IFNγ and IL-2 producing CD4^+^ T-cells were measured in a subgroup of patients using commercially available kits (IFNγ Elispot (Diaclone, Besançon France) and IL-2 Fluorospot (C.T.L Europe, Bonn, Germany). For more detailed description see Supplementary Methods.

### Immunogenicity assessment

The primary endpoint was the proportion of subjects with an increase rate in neutralizing antibody titers of at least 10-fold, measured by a microneutralization technique, between day 0 and day 15 against D614 SARS-CoV-2 viral strain or B.1.351 variant.

The main other prespecified immunological endpoints were the rate of increase between day 0 and day 15 in neutralizing antibody titers against SARS-CoV-2 Wuhan (D614) and variants Beta, Delta and Omicron BA.1, geometric mean of anti-Spike IgG levels (expressed as BAU/mL) and IFNγ and IL-2 secreting CD4^+^ T-cells after stimulation with Spike peptides derived from wild-type SARS-CoV-2 or Omicron variant in each randomized group.

### Safety assessment

Injection-site and systemic solicited adverse events were collected for 7 days and unsolicited adverse events through 28 days after vaccination using diary cards provided to each participant. In the grading of adverse events, the FDA Toxicity Grading Scale (2007) was used (from grade 0, sign/symptom within normal limits to 4, life-threatening adverse event) for the following solicited adverse event: pain, arthralgia, asthenia or malaise, headache, fever, chills, swelling, lymphadenopathy, myalgia, nausea, edema, redness, vomiting; the WHO scale (mild, moderate or severe) was used for the following solicited adverse events, not present in FDA scale: itching, diarrhea, pain in extremities, insomnia.

### Statistical analysis

The intent-to-treat (ITT) population included all randomized participants except those with positive nucleoprotein antibody serology at inclusion. Per protocol (PP) population included all randomized vaccinated participants without major protocol deviations, except participants presenting a positive nucleoprotein serology at day 0 or day 15, SARS CoV-2 infection after boost and lost to follow-up participants. The safety population included all randomized participants who received the vaccine booster dose. The immunogenicity analysis was performed on the per-protocol population.

As no data was available on the BNT162b2 vaccine, the sample size calculation was based on published data on the mRNA-1273 vaccine in which an increase rate of neutralizing antibody titer of 23 against ancestral SARS-CoV-2 (D614G) and 32 for the B.1.351 variant after mRNA-1273 boost was described.^13^ Using a conservative approach, we considered neutralizing activity to be sufficient if the increase rate was at least 10 at D15. We assumed a proportion of subjects with an increase rate of at least 10 between D0 and D15 of 80%. One hundred subjects per group allowed an estimation of this proportion with a 95% confidence interval (CI) of 7.8%. Thus, a total of 300 volunteers had to be randomized (100 per group).

For each viral strain (D614 and B.1.351), the primary endpoint was described using frequencies, percentages and 95% CIs calculated using the exact method. In a post hoc analysis, groups were compared using Chi-2 test.

Baseline patient characteristics were described overall and for each group using the number (percentage) for categorical variables and the mean (SD) or median [interquartile range], according to distribution, and range for quantitative variables. For each group, the anti-SARS-CoV-2 IgG antibody titers directed against the S1 domain of the spike protein and the neutralizing antibody titers measured by a microneutralization technique against Wuhan strain (D614) and variants (B.1.351, Delta, Omicron BA.1) measured at day 0, day 15 were described as geometric means with two-sided 95% confidence intervals (95% CI). Adverse reactions and events were described using frequencies and percentages.

The statistical analysis was conducted using SAS software version 9.4 (SAS Institute, Inc., Cary, North Carolina, USA). R freeware (version 3.6.3) and GraphPad Prism software (version 9.2.0, San Diego, California USA) were used for the graphs.

## RESULTS

### Study participants

A total of 247 participants who had received two doses of BNT162b2 were randomized from December 8, 2021 to January 14, 2022 to receive a third dose: 85 in the group Sanofi/GSK MV(D614), 80 in the group Sanofi/GSK MV(Beta) and 82 in the group BNT162b2. Details on causes for exclusion from immunogenicity analysis are presented in **Figure 1**. Because of the start of the study after the beginning of booster vaccination campaign in elderly, the enrollment of subjects over 65 years of age was difficult, and the number of inclusion of those under 65 years was increased. The enrollment was planned to stop no later than January 14.

**Figure 1.**
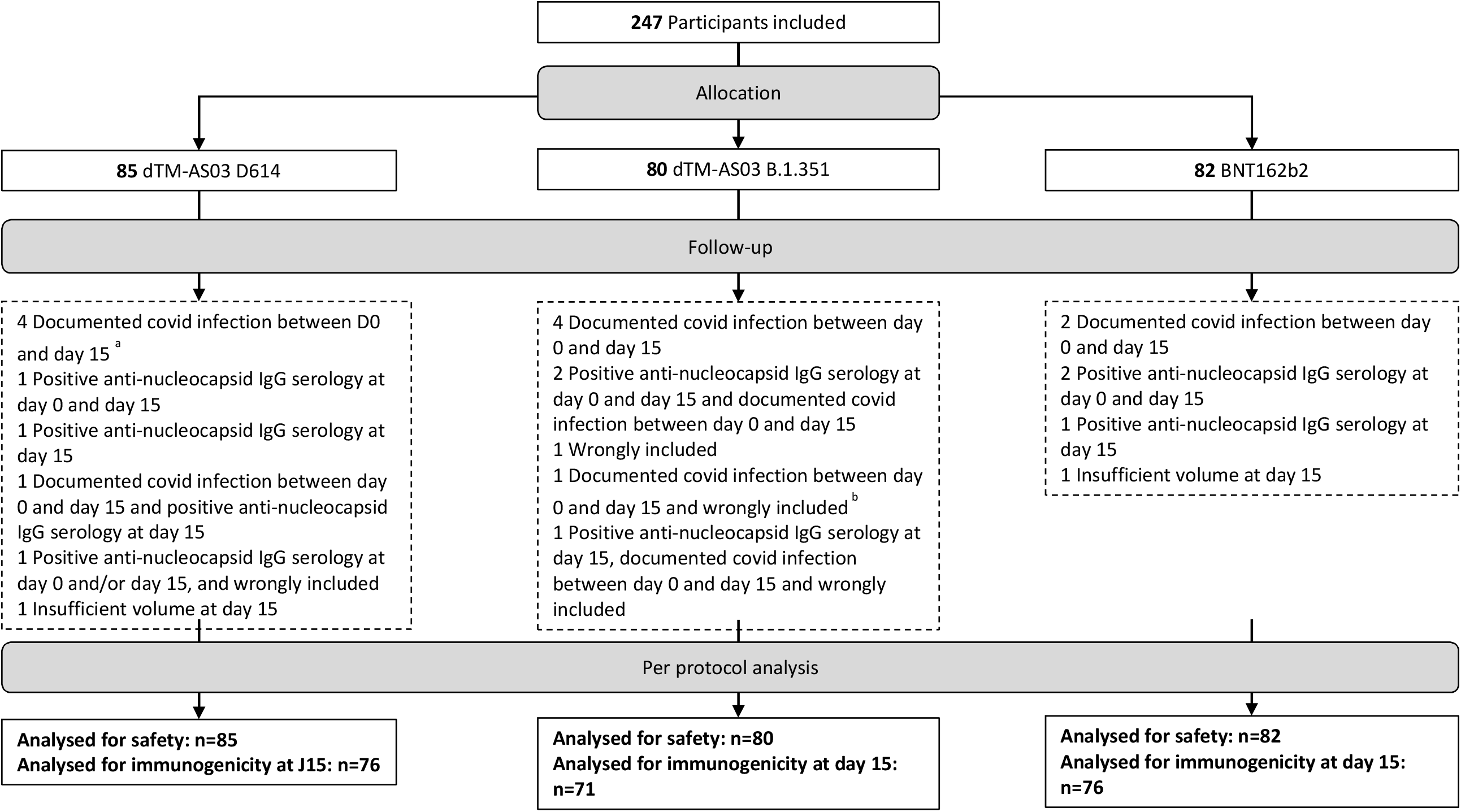
Flow chart. ^a^ Day 15 visit and blood sample not performed ^b^ Day 15 visit performed by phone and blood sample not performed

The mean age of participants in the per-protocol population was 40.6 years and the proportion of women was 40.4%. The baseline characteristics of the three randomized groups were comparable (**Table 1**). Infection by SARS-CoV-2 between day 0 and day 15 or positive SARS-CoV-2 anti-nucleocapsid IgG serology at day 0 or day 15 were reported in 8.5% (21/247) of patients (n=8 [9.4%] in the group Sanofi/GSK-D614; n=8 [10.0%] in the group Sanofi/GSK-B.1.351; n=5 [6.1%] in the group Pfizer BNT162b2). The per-protocol population for immunogenicity included 223 participants: 76 in the group Sanofi/GSK MV(D614), 71 in the group Sanofi/GSK MV(Beta) and 76 in the group Pfizer BNT162b2.

**Table 1.** Characteristics of patients at inclusion (per-protocol population).

|  | **Total population**<br>**(n=223)** | **Sanofi/GSK-**<br>**D614**<br>**(n=76)** | **Sanofi/GSK-**<br>**B.1.351**<br>**(n=71)** | **Pfizer BNT162b2**<br>**(n=76)** |
| --- | --- | --- | --- | --- |
| Age, y |  |  |  |  |
| Mean (SD) | 40.6 (13.0) | 40.2 (13.5) | 41.4 (11.3) | 40.4 (13.9) |
| Range | 18–73 | 18–73 | 22–68 | 20–69 |
| Female gender, n (%) | 90 (40.4) | 29 (38.2) | 23 (35.2) | 36 (47.4) |
| Body mass index, kg/m<sup>2</sup> |  |  |  |  |
| Mean (SD) | 25.0 (4.5) | 25.2 (4.5) | 25.4 (4.5) | 24.4 (4.6) |
| Range | 15.2–40.8 | 16.8–35.6 | 18.5–40.8 | 15.2–40.4 |
| Current smoker, n (%) | 49 (22.1) | 16 (21.3) | 17 (23.9) | 16 (21.1) |
| Comorbidity, n (%) |  |  |  |  |
| Obesity <sup>a</sup> | 27 (12.1) | 13 (17.1) | 8 (11.3) | 6 (7.9) |
| Hypertension | 11 (4.9) | 6 (7.9) | 2 (2.8) | 3 (3.9) |
| Dyslipidemia) | 6 (2.7) | 4 (5.3) | 1 (1.4) | 1 (1.3) |
| Diabetes | 2 (0.9) | 2 (2.6) | 0 | 0 |
| Time between 1<sup>st</sup> and 2<sup>nd</sup> dose, days |  |  |  |  |
| Median (IQR) | 39 (31; 42) | 39 (30; 42) | 39 (33; 42) | 38 (32; 40) |
| Range | 21–49 | 21–49 | 21–44 | 21–42 |
| Time between 2<sup>nd</sup> and 3<sup>rd</sup> dose, days |  |  |  |  |
| Median (IQR) | 174 (164; 187) | 176 (167.5; 188) | 171 (164; 184) | 174.5 (160; 188) |
| Range | 121–223 | 121–211 | 148–223 | 141–212 |

<sup>a</sup> Body mass index > 30 kg/m<sup>2</sup>

### Humoral immune response

The proportion of participants with at least a 10-fold increase of neutralizing antibody titers between day 0 and day 15 for the Wuhan (D614) SARS-CoV-2 strain were 55.3% (95% CI 43.4–66.7) for Sanofi/GSK MV(D614), 76.1% (95% CI 64.5–85.4) for Sanofi/GSK MV(Beta) and 63.2% (95% CI 51.3–73.9) for Pfizer BNT162b2 (p=0.03). For the B.1.351 (Beta) viral strain, these rates were 44.7% (95% CI 33.3–56.6), 84.5% (95% CI 74.0–92.0) and 51.3% (95% CI 39.6–63.0), respectively (p<0.0001) (**Supplementary Table 1**).

The geometric mean titers of the neutralizing against Wuhan (D614) strain and Beta, Delta and Omicron BA.1 variants in the three randomized groups are presented in **Figure 2** and **Supplementary Table 2**. The increase of the titers of neutralizing antibodies was higher in patients whose neutralizing antibodies were below the threshold of positivity at day 0.

**Figure 2.**
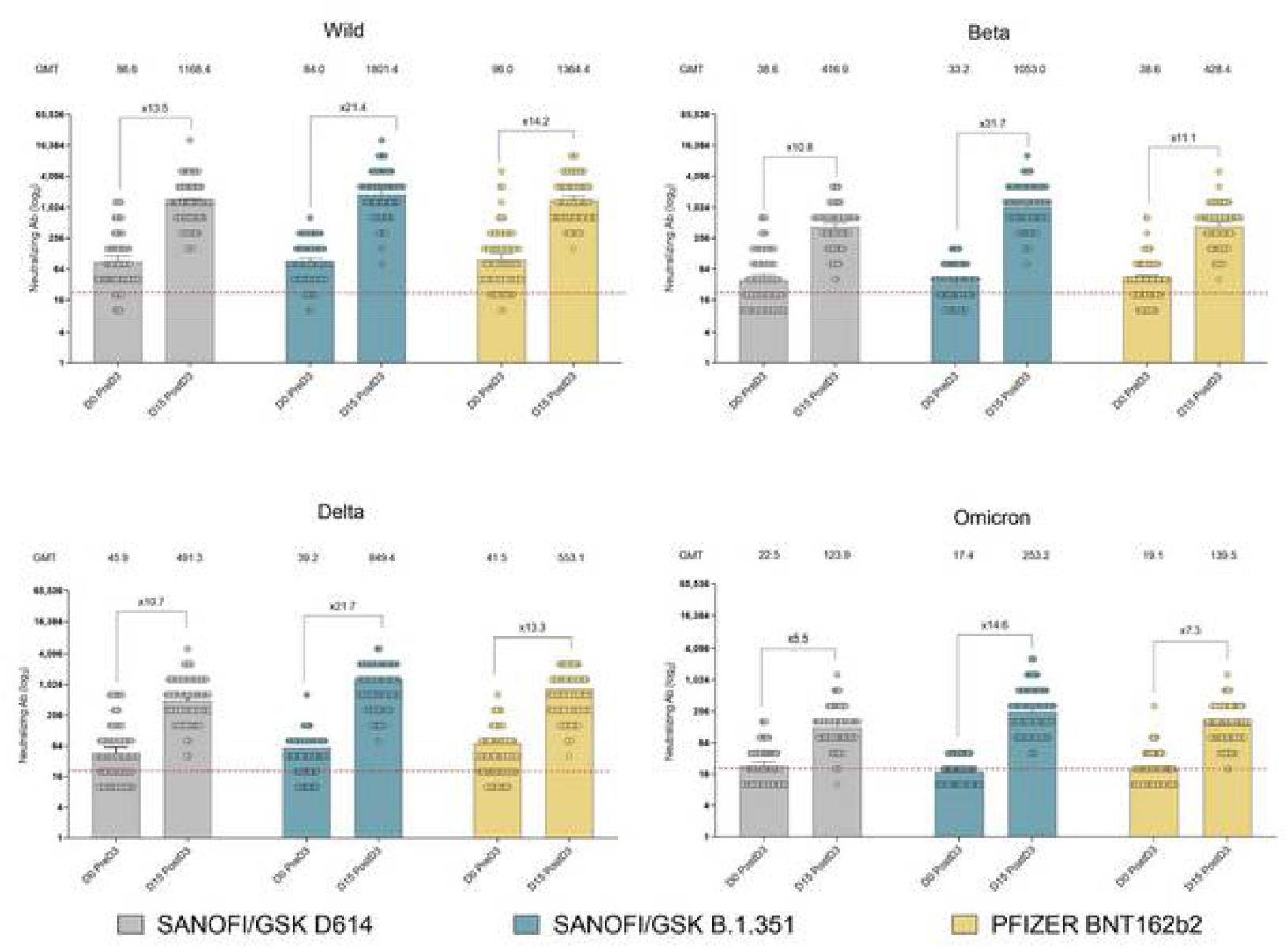
Neutralizing antibodies against D614 (wild-type; Wuhan) SARS-CoV-2 and variants Beta, Delta and Omicron BA.1 at D0 andD15 after the boost dose (“post D3”)with Sanofi/GSK-D614, Sanofi/GSK-B.1.351 or BNT162b2 (per-protocol population); dotted line represents the positivity threshold.

The geometric mean titer of anti-S1 increased from 277.1 at day 0 to 1875.1 BAU/mL at day 15 in Sanofi-GSK MV(D614) group, from 206.8 to 2240.8 BAU/mL in Sanofi-GSK MV(Beta) group and from 253.6 to 2405.4 BAU/mL in BNT162b2 group (**Supplementary Table 3**).

### Cellular immune responses

IFNγ and IL-2 producing CD4+ T-cells were increased after vaccination when T cells were stimulated with Spike peptides derived from wild-type SARS-CoV-2; pool of peptides derived from the Omicron variant also enhance IFNγ secreting CD4^+^ T-cells in the three randomized groups (**Figure 3**). The rates of CD4^+^ T-cells with positive vaccine induced -response at day 15 for IL-2 and IFNγ/IL-2 were significantly increased in patients boosted by Sanofi/GSK MV(Beta) compared to BNT162b2 after stimulation with Spike peptides of wild-type SARS-CoV-2 (**Supplementary Figure 1**).

**Figure 3.**
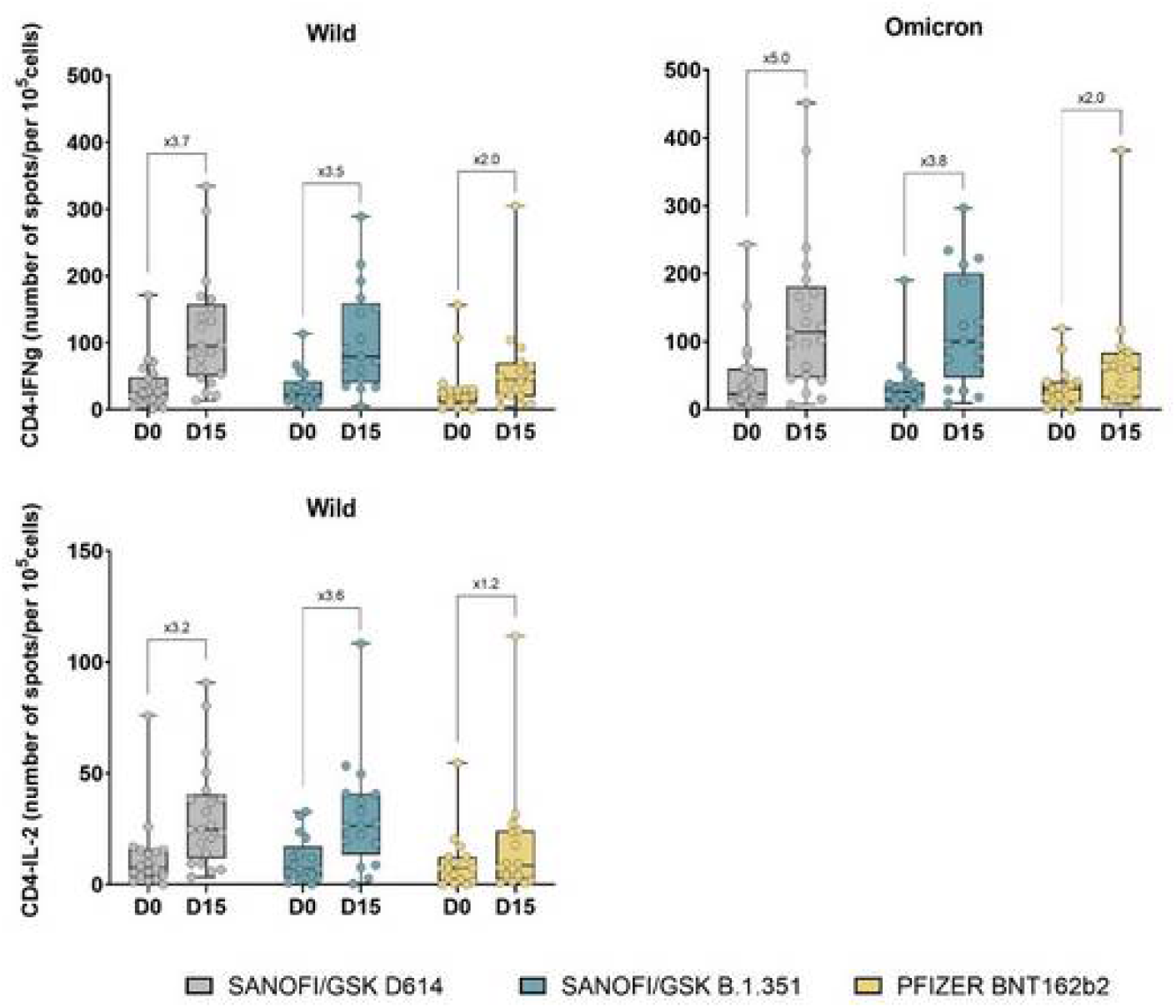
Induction of TH1-CD4^+^ T-cell response after a boost with BNT162b2 mRNA or Sanofi/GSK D614 or Sanofi/GSK B.1.351 vaccines. CD4^+^ T-cells were purified from PBMC collected before or 15 days after boosting by the various vaccines. IFNγ Elispot and IL-2 Fluorospot assays were performed by incubating CD4^+^ T-cells with a pool of 15-mer overlapping peptides derived from the wild type (Wuhan) SARS-CoV-2 for both assays or from the Omicron variant of SARS-CoV-2 for the detection of IFNg producing cells. The number of spots was enumerated on an Elispot/Fluorospot reader with a positivity threshold set up at 10 spots per 10^5^ cells. The increase between the medians at D15 and D0 for the various CD4^+^ T-cell population is shown.

### Vaccine safety

Solicited injection-site adverse events from day 0 to day 7 were common and reported by 83.5% of participants in Sanofi/GSK MV(D614) group, 80.0% in Sanofi/GSK MV(Beta) group and 82.9% in BNT162b2 group (**Figure 4** and **Supplementary Table 4**). The most frequent injection-site adverse event was pain (83.5%, 77.5% and 80.5%, respectively). Solicited systemic adverse event from D0 to D7 were reported by 48.2% of participants in Sanofi/GSK D614 group, 62.5% in Sanofi/GSK B.1.351 group and 62.2% in BNT162b2 group. The most frequent systemic adverse events were asthenia/malaise (31.8%, 40.0% and 35.4%, respectively) and headache (27.1%, 33.7% and 42.7%). The rates patients who reported injection-site or systemic adverse events were comparable between the three randomized groups; their severity was mainly Grade 0–2.

**Figure 4.**
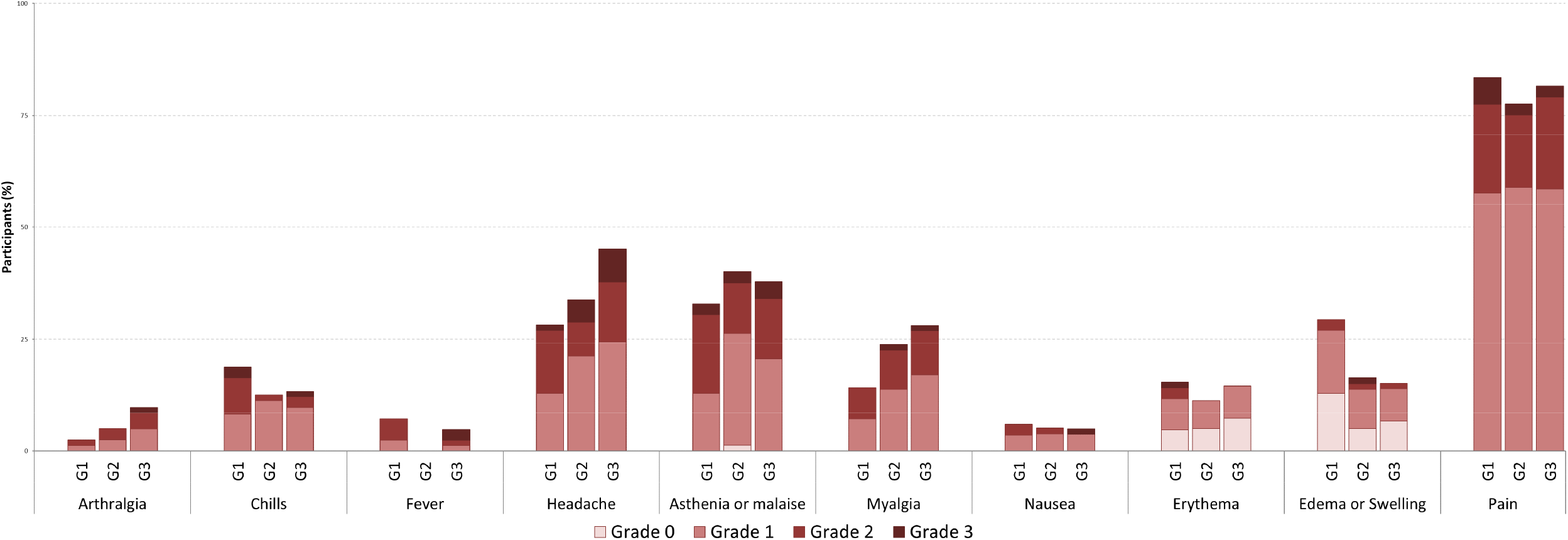
Rates and grades of severity of solicited adverse events reported from D0 to D7 by participants from the three randomized groups of the safety population (G1, Sanofi/GSK-D614; G2, Sanofi/GSK-B.1.351; G3, BNT162b2) according to the Toxicity Grading Scale for Healthy Adult and Adolescent Volunteers Enrolled in Preventive Vaccine Clinical Trials (Modified FDA scale/September 2007).

Two serious adverse events were reported. Sanofi-GSK MV(Beta): severe polyarthritis flare-up 3 days after vaccination in a woman in her 60s with known rheumatoid arthritis with previous flare-ups during the first mRNA vaccine injections (considered related to the vaccine; the Data and Safety Monitoring Board considered that the patient was wrongly enrolled even if the patient was not receiving immunosuppressive therapy at the time of inclusion). Sanofi/GSK MV(D614): appendectomy in a man in his 30s 15 days after vaccine administration (possible relationship). Both serious adverse events resolved without sequalae.

## DISCUSSION

In this randomized, single-blinded, multicenter trial, both homologous (with mRNA BNT162b2) and heterologous (with D614 and B.1.351 Sanofi/GSK adjuvanted recombinant protein vaccines and AS03 as adjuvant) boost immunizations were well tolerated and immunogenic in patients primed with two doses of BNT162b2 vaccine both against the ancestral Wuhan (D614) SARS-CoV-2 strain, B.1.351 (Beta) variant and recent circulating variants of concern (VoC). A higher percentage of subjects with a ≥10-fold increase rate in neutralizing antibody titers 15 days after the booster dose was observed with the Sanofi/GSK MV(Beta) vaccine against SARS-CoV-2 D614 and B.1.351 viral strains. The three vaccines also elicited neutralizing antibodies against Delta and Omicron variants, with higher response with Sanofi/GSK MV(Beta) vaccine.

Several studies have compared homologous and heterologous third dose (booster) following two doses of COVID-19 vaccines. The study of Munro *et al* (COV-BOOST) performed in United Kingdom assessed seven different COVID-19 vaccines as a third dose after two doses of ChAdOx1 (AstraZeneca) or BNT162b2.^14^ All vaccines boosted neutralizing antibodies after ChAdOx1 primo-vaccination and all except one after BNT162b2 primo-vaccination (the Valneva vaccine did not achieve the predefined criteria). In the Phase I-II study of Atmar *et al* performed in the United States, adults who had completed a COVID-19 vaccine regimen with mRNA1273 (Moderna), Ad26.COV2.S (Johnson & Johnson-Janssen) or BNT162b2 received a booster injection with one of the three vaccines.^15^ Heterologous boosting provided comparable or higher levels of immunogenicity levels. Sableroles *et al* assessed the immunogenicity of three different vaccine boosters (Ad26.COV2.S, mRNA-1273 or BNT162b2) or no boost after Ad26.COV2.S priming.^16^ All three vaccines were immunogenic, but the strongest responses were observed with heterologous boosting (*i*.*e*., mRNA vaccines), more specifically with mRNA-1273. The study of Barros-Martins assessed the heterologous prime-boost vaccination with ChAdOx1-Cov-19/BNT162b2 compared to the homologous vaccination with ChAdOx1-Cov-19.^5^ Although both regimens boosted prime-induced immunity, heterologous boosting with BNT162b2 elicited significantly higher titers of neutralizing antibodies, particularly against the B.1.1.7 (Alpha), B.1.351 (Beta) and P.1 (Gamma) variants. Comparable results were reported by Schmidt *et al* in a German study in participants primed with ChAdOx1-Cov-19 vaccine and then boosted with mRNA vaccines (BNT162b2 or mRNA-1273).^6^ A Chinese study compared the heterologous boosting with the recombinant adenovirus type 5-vectored COVID-19 vaccine Convidecia (AD5-nCOV) to the homologous boosting with Sinovac COVID-19 vaccine (CoronaVac; inactivated virus COVID-19 vaccine).^17^ Heterologous boosting with Convidecia elicited significantly higher rates of neutralizing antibodies than homologous boosting with CoronaVac. Taken together these results suggest that immunogenicity after heterologous prime-boost leads to comparable or even higher immune response than homologous prime-boost.

These data support our finding of higher immunogenicity induced by a heterologous priming-boost with BNT162b2 followed by the Sanofi/GSK adjuvanted recombinant protein vaccine MV(Beta) than the homologous prime-boost with Pfizer BNT162b2. In addition, we showed that the B.1.351 variant (Beta) formulation of the new Sanofi/GSK vaccine provided better cross-neutralization response against both the ancestral strain but also variants such as Delta and Omicron BA.1 compared with the mRNA-based BNT162b2 and also with the Sanofi/GSK MV(D614) vaccine. This same formulation also increased the rate of vaccine-induced double positive producing IFNγ and IL-2 or IL-2 CD4^+^ T-cell response (**Supplementary Fig 1**). In general, the ability of CD4^+^ T-cells to produce IL-2 or multiple cytokines is associated with their long-term survival and better pathogen protection.^18,19^ In HIV infection, viral controllers had significantly more CD4^+^ T-cells co-expressing IL-2 and IFNγ than other HIV patient categories.^20^

This study has some limitations. Compared to the at-risk population for severe forms of SARS-CoV-2 infection, the study population was younger and included a smaller percentage of people ≥65 years than previously planned. Indeed, by the time the study was started, the elderly population had already received a third dose. Another limitation is the priming with a unique vaccine. The primary endpoint was based on an increase in neutralizing antibodies against the Wuhan (D614) and B.1.351 (Beta) strains, which are variants of SARS-CoV-2 that no longer circulate. However neutralizing antibodies against more recent variants were also evidenced.

Despite these limitations, our study is the first to report immunization and reactogenicity data on the heterologous boost with an adjuvanted recombinant protein vaccine containing a variant of concern different from the one present at the priming. The higher neutralizing response elicited by the vaccine containing the Beta Spike protein will need further investigations to understand the mechanisms underlying these results. Of interest, this higher immunogenicity was not associated with higher reactogenicity. However, the observed higher immunogenicity should be interpreted with caution, as the relationship between antibody levels at 15 days and the long-term protection remains to be fully characterized.

These results, which address the possibility of combining different vaccines for priming and boosting, are important for vaccination campaigns and for improving vaccination coverage in countries that are still under-vaccinated. Indeed, recent vaccines or those still under development may have some advantages for the implementation of booster vaccination, such as improved immunogenicity, reactogenicity, availability, thermostability and potentially better acceptability for certain populations.

In conclusion, all three vaccines boosted neutralizing antibodies after BNT162b2 initial course with no safety concerns. Heterologous boosting with the Sanofi/GSK Beta formulation vaccine resulted in a higher neutralizing antibody response against Beta variant but also the original strain and Delta and Omicron BA.1 variants, compared with mRNA BNT162b2 vaccine or the Sanofi/GSK MVD614 formulation. New vaccines containing Beta spike protein may represent an interesting strategy for broader protection against SARS CoV-2 variants.

## Data Availability

All data produced in the present work are contained in the manuscript

## Contributors

OL, AR, XDL, ET, LB and TS designed the study. AT, LC, OL, EK, MC, LBL, LN, ML, IBG, MB, CSM, RF, KL, LS, FL, JSA, EBN, MPT, CC, PP, BD, SP, DD, NBH and AB acquired, analysed or interpreted the study. OL, MC, EK, AR, LN, XDL, ET, LBL, ML, IBG, MB, CSM, RF, KL, LS, FL, JSA, EBN, MPT, CC, PP, BD, SP and DD drafted the manuscript. OL, TS, AR, XDL, MC and ET critically reviewed the manuscript for important intellectual content. MC and AR did the statistical analysis. OL obtained funding. CP, AT, LC EK, EN, FC, NBH and AB were in charge of administrative, technical and material support. OL and TS supervised the study. AB, PL, EL and LW constituted the independent monitoring committee. TS and OL had full access to all of the data in the study and takes responsibility for the integrity of the data and the accuracy of the data analysis.

## Declaration of interest

OL reports grant from French ministry of health, grant and some personnal fees for board membership from Pfizer, Sanofi Pasteur, MSD, GSK, CJ reports fees for boardmembership from Astrazeneca, Pfizer, MSD. TS reports grant from DGOS (French Ministry of health), Asatrazeneca, Bayer, Novartis, Sanofi, Boehringer, Daiichi Sankyo, Eli Lilly, GSK and personal fees for boardmembership and consulting from various pharmaceutical companies, including Astrazeneca, Sanofi, Bayer, Ablative Solutions and 4 Living Biotech. KL reports personal fees and Grant/Research Support from Gilead, MSD, Janssen, ViiV, Spikimm and fees for development of educational presentations from Janssen, Gilead, MSD, Sobi, and Chiesi. EBN has received grant pending from Sanofi Pasteur and fees for board membership from Pfizer, and Janssen. LBLN received personal fees for advisory experts and participation to conference from Pfizer. XDL reports research grant from INSERM.

## Funding/Support

This study was supported by the French Ministry of Solidarity and Health and Sanofi and sponsored by Assistance Publique HÉpitaux de Paris.

## Independent monitoring committee

Brigitte Autran, Edouard Lhomme, Paul Loubet and Laurence Weiss.

## COVIBOOST Study Group

Bertrand Accart, Lauren Agnelli, Olivia Aranda, Lucile Armelout, Laurence Attolini, Marc Bardou, Sophie Bayer, Narimane Benhamadi, Virginie Bernigal, Jeanne Bertona, Olivier Blin, Alexandre Bolle, Camille Bourgneuf, Corinne Brochier, Serge Bureau, Christine Caloustian, Estelle Canton, Nadine Casimir, Julie Charligny, Edith Chaze, Anne Conrad, Charlotte Cosse, Virginie Costenoble, AmÇlie Cransac, DÇborah Dauvet, Juliette De Langhe, Maelle Detoc, Odaliz Duverneil, Elise Elrezzi, Soumaya Essalim, Kahina Fali, Jose Fernandes, Pascale Fouilloux, ValÇrie Galvan, Akila Ghezli, Nicolas Gonnet, Corinne Guerin, Audrey Guillie, Marie Gyselinck, Nacilla Haicheur, Lamia Hamoudi, Michael Hisbergues, Anne Hutt-Clauss, Vinca Icard, Julie Jambon, Rafik Kafi, Marie-Aude Kamp, Emmanuelle Kasprzyk, Laure Lalande, Marie-Noélle Lefebvre, Marc Leone, Estelle Letienne, Benjamin Leveau, Emmanuelle Liegey, Maxime Logier, Maxime Luu, Cecilia Mallmann, LÇa Marchand, Julie MarÇchal, Jacob Marion, Laurent Martin, Maria-Claire Migaud, Cecile Mignon, Jade Montlahuc, Sophie Morange, Alexia Moulin, Stanislas Ogoudjobii, Gaélle Pagand, CÇline Pago, BÇatrice Parfait, Elodie Perrot, Florian Prever, Julie Quantin, Muriel Quillard, Patrick Rasoamanana, Helene RÇnaux, Estelle Revy, NoÇmie Roberjot, Laurence Rodari, Franck Rouby, LorÅne Roux, Karen Sagorny, Betty Schoemaecker, StÇphanie Somers, Katya Touat, Mamadouba Toure, David VallÇe, Marie Vayssettes, Françoise Vignaud, Jean-Louis Vincent, Nawal Waucquier, Yu Jin Yung.

## Notes

### Competing Interest Statement

Odile Launay 
- AstraZeneca (Financial)
- GlaxoSmithKline (Advisor/Consultant, Grant/Research Support)
- Johnson & Johnson (Advisor/Consultant, 
Grant/Research Support)
- MD (Advisor/Consultant)
- Moderna (Advisor/Consultant)
- MSD (Data safety monitoring board)
- Pfizer (Advisor/Consultant, Grant/Research Support)
- Sanofi Pasteur (Advisor/Consultant, Grant/Research Support, Data safety monitoring board)
Tabassome Simone: 
-DGOS (French Ministry of health),  Astrazeneca, Bayer, Novartis, Sanofi, Boehringer, Daiichi Sankyo, Eli Lilly, GSK:  grant 
-Astrazeneca, Sanofi, Bayer, Ablative Solutions and 4 Living Biotech: personal fees for boardmembership and consulting  
Karine Lacombe:
- Gilead, MSD, Janssen, ViiV, Spikimm (fees and Grant/Research Support) 
-Janssen, Gilead, MSD, Sobi, and Chiesi  (fees for development of educational presentations). 
Elisabeth Botelho-Nevers
-Sanofi Pasteur (grant pending from) 
- Pfizer and Janssen (fees for board membership) 
Liem Binh Luong Nguyen: 
Pfizer (personal fees for advisory experts and participation to conference). 
Xavier De Lamballerie/
-INSERM (research grant).

### Clinical Trial

NCT05124171

### Funding Statement

This study was funded by the French Ministry of Solidarity and Health and Sanofi

### Author Declarations

Ethics Committee/IRB of Comite de Protection des Personnes Ile de France III and the French Health Products Safety Agency gave ethical approval for this work

