## Supplemental data for "Immunogenicity and Safety of Beta Adjuvanted Recombinant Booster Vaccine"

### SUPPLEMENTARY DATA

#### Spike-specific IFNγ or IL-2 producing CD4$^+$ T-Cells

The cellular immune response was assessed *in vitro* by measuring production of IFNγ by CD4+ T-cells by ELISPOT (Diaclone) and IL-2 secreting CD4$^+$ T-cells by FLUOROSPOT (C.T.L) after stimulation with a pool of 15-mer overlapping peptides derived from the wild type (Wuhan) SARS-CoV-2 for both assays or from the Omicron variant of SARS-CoV-2 only for IFNγ Elispot, at concentration of 1 µg/ml. The samples tested were collected at baseline (D0) and D15 after the vaccine boost. CD4+ T-cells were obtained from defrost PBMC by a positive selection with a system MACS cell Separation using beads CD4 and LD columns (Miltenyi Biotec, Paris). CD4$^+$ T-cells were sensitized with the pool of spike peptides derived from the wild type SARS-CoV-2 Wuhan strain (JPT peptide technologies, Berlin, Germany) or the Omicron variant (peptides&elephants, Hennigsdorf, Germany) or a negative control (unstimulated cells in CTL-test medium, Bonn, Germany) or a positive control (cells stimulated with PMA-ionomycin (Sigma Saint-Quentin-Fallavier, France) for 20 h in a cell incubator at 37°C. After incubation, plates were revealed according manufacturer instructions, then scanned and analyzed on a C.T.L reader (S6 Ultimate). A response was considered positive if the number of spots in the wells stimulated with the spike specific peptides was twofold higher than the number of spots in the negative control using a cutoff of 10 SFC/$10^5$ cells after background subtraction as previously detailed (Sirima *et al*, *Lancet Infect Dis*. 2020;20:585-597). A positive response to vaccine was set up as a two-fold increase of the cytokine producing spike specific CD4$^+$ T-cell at D15 compared to D0.

**Supplementary Table 1.** Rates of patient with at least a 10-fold increase of neutralizing antibodies against SARS-CoV-2 D614 and B.1.351 viral strains between D0 and D15 (per-protocol and intent-to-treat populations).

| | **Sanofi/GSK- D614** | **Sanofi/GSK- B.1.351** | **Pfizer BNT162b2** | **P-value** |
|---|---|---|---|---|
| **Neutralizing antibodies (increase ≥ 10-fold)** | | | | |
| **Per-protocol population** | **(N=76)** | **(N=71)** | **(N=76)** | |
| Wuhan (D614) viral strain, n (%) | 42 (55.3) | 54 (76.1) | 48 (63.2) | 0.030 |
| 95% CI, % | 43.4–66.7 | 64.5–85.4 | 51.3–73.9 | |
| B.1.351 (Beta) viral strain, n (%) | 34 (44.7) | 60 (84.5) | 39 (51.3) | < 0.0001 |
| 95% CI, % | 33.3–56.6 | 74.0–92.0 | 39.6–63.0 | |
| **Intent-to-treat population** | **(N=84)** | **(N=78)** | **(N=80)** | |
| Wuhan (D614) viral strain, n (%) | 48 (57.1) | 61 (78.2) | 51 (63.8) | 0.016 |
| 95% CI, % | 45.9–67.9 | 67.4–86.8 | 52.2–75.2 | |
| B.1.351 (Beta) viral strain, n (%) | 41 (48.8) | 66 (84.6) | 41 (51.3) | < 0.0001 |
| 95% CI, % | 37.7–60.0 | 74.7–91.8 | 51.3–62.6 | |

**Supplementary Table 2.** Neutralizing antibodies against D614 (wild type; Wuhan) SARS-CoV-2 and Beta, Delta and Omicron BA.1 variants in the three randomized groups (per-protocol population).

| | Sanofi/GSK-D614 (N=76) | | Sanofi/GSK-B.1.351 (N=71) | | Pfizer BNT162b2 (N=76) | |
|---|---|---|---|---|---|---|
| | **N[a]** | **GMT (95% CI)** | **N[a]** | **GMT (95% CI)** | **N[a]** | **GMT (95% CI)** |
| Wild-type (D614; Wuhan) | | | | | | |
| D0 | 76 | 86.8 (66.0; 114.3) | 71 | 84.0 (67.5; 104.6) | 76 | 96.0 (71.7; 128.6) |
| D15 | 76 | 1168.4 (928.0; 1471.2) | 71 | 1801.4 (1414.8; 2293.6) | 76 | 1364.4 (1091.9; 1704.8) |
| Beta variant | | | | | | |
| D0 | 76 | 38.6 (29.4; 50.5) | 71 | 33.2 (27.5; 40.2) | 76 | 38.6 (31.2; 47.7) |
| D15 | 76 | 416.9 (334.2; 520.0) | 71 | 1053.0 (840.3; 1319.5) | 76 | 428.4 (346.6; 529.7) |
| Delta variant | | | | | | |
| D0 | 76 | 45.9 (34.7; 60.6) | 71 | 39.2 (32.0; 48.1) | 76 | 41.5 (33.1; 52.0) |
| D15 | 76 | 491.3 (393.3; 613.7) | 71 | 894.4 (678.6; 1063.3) | 76 | 553.1 (443.2; 690.2) |
| Omicron BA.1 variant | | | | | | |
| D0 | 76 | 22.5 (18.8; 27.0) | 71 | 17.4 (15.3; 19.9) | 76 | 19.1 (16.3; 22.4) |
| D15 | 76 | 123.9 (101.5; 151.4) | 71 | 253.2 (201.8; 317.6) | 76 | 139.5 (115.0; 169.3) |

[a] Available data.

**Supplementary Table 3.** Anti-S1 antibodies from day 0 to day 15 (per-protocol population).

| | Sanofi/GSK-D614 (N=76) | | Sanofi/GSK-B.1.351 (N=71) | | Pfizer BNT162b2 (N=76) | |
|---|---|---|---|---|---|---|
| | N[a] | GMT (95% CI) | N[a] | GMT (95% CI) | N[a] | GMT (95% CI) |
| **Anti-S1 antibodies, BAU/mL, GMT (95% CI)** | | | | | | |
| Day 0 | 76 | 277.1 (229.7; à 334.3) | 71 | 206.8 (134.5; 318.0) | 76 | 253.6 (214.4; 300.0) |
| Day 15 | 76 | 1875.1 (1628.0; 2159.6) | 71 | 2240.8 (1931.3; 2600.0) | 76 | 2405.4 (2130.8; 2715.4) |

**Supplementary Table 4.** Solicited injection-site and systemic adverse events (AE) in safety population.

| | SANOFI/GSK-D614 n=85 | SANOFI/GSK-B.1.351 n=80 | PFIZER BNT162b2 n=82 |
|---|---|---|---|
| | | n (%) | |
| **At least one solicited AE between D0 and D7** | 77 (90.6) | 71 (88.8) | 75 (91.5) |
| **At least one injection-site AE between D0 and D7** | 71 (83.5) | 64 (80.0) | 69 (84.1) |
| **Pain** | | | |
| No | 14 (16.5) | 18 (22.5) | 15 (18.3) |
| Grade 1 | 49 (57.6) | 47 (58.8) | 48 (58.5) |
| Grade 2 | 17 (20.0) | 13 (16.3) | 17 (20.7) |
| Grade 3 | 5 (5.9) | 2 (2.5) | 2 (2.4) |
| **Redness** | | | |
| No | 72 (84.7) | 71 (88.8) | 70 (85.4) |
| Grade 0 | 4 (4.7) | 4 (5.0) | 6 (7.3) |
| Grade 1 | 6 (7.1) | 5 (6.3) | 6 (7.3) |
| Grade 2 | 2 (2.4) | 0 (0) | 0 (0) |
| Grade 3 | 1 (1.2) | 0 (0) | 0 (0) |
| **Edema or swelling** | | | |
| No | 60 (70.6) | 67 (83.8) | 69 (84.1) |
| Grade 0 | 11 (12.9) | 4 (5.0) | 6 (7.3) |
| Grade1 | 12 (14.1) | 7 (8.8) | 6 (7.3) |
| Grade 2 | 2 (2.4) | 1 (1.3) | 1 (1.2) |
| Grade 3 | 0 (0) | 1 (1.3) | 0 (0) |
| **Itching** | | | |
| No | 77 (90.6) | 75 (93.8) | 78 (95.1) |
| Mild | 8 (9.4) | 5 (6.3) | 3 (3.7) |
| Moderate | 0 (0) | 0 (0) | 1 (1.2) |
| **At least one systemic AE between D0 and D7** | 42 (49.4) | 50 (62.5) | 53 (64.6) |
| **Asthenia or malaise** | | | |
| No | 57 (67.1) | 48 (60.0) | 51 (62.2) |
| Grade 0 | 0 (0) | 1 (1.3) | 0 (0) |

| | SANOFI/GSK-D614 n=85 | SANOFI/GSK-B.1.351 n=80 | PFIZER BNT162b2 n=82 |
|---|---|---|---|
| | | n (%) | |
| Grade 1 | 11 (12.9) | 20 (25.0) | 17 (20.7) |
| Grade 2 | 15 (17.6) | 9 (11.3) | 11 (13.4) |
| Grade 3 | 2 (2.4) | 2 (2.5) | 3 (3.7) |
| **Arthralgia** | | | |
| No | 83 (97.6) | 76 (95.0) | 74 (90.2) |
| Grade 1 | 1 (1.2) | 2 (2.5) | 4 (4.9) |
| Grade 2 | 1 (1.2) | 2 (2.5) | 3 (3.7) |
| Grade 3 | 0 (0) | 0 (0) | 1 (1.2) |
| **Headache** | | | |
| No | 61 (71.8) | 53 (66.3) | 45 (54.9) |
| Grade 1 | 11 (12.9) | 17 (21.3) | 20 (24.4) |
| Grade 2 | 12 (14.1) | 6 (7.5) | 11 (13.4) |
| Grade 3 | 1 (1.2) | 4 (5.0) | 6 (7.3) |
| **Fever** | | | |
| No | 79 (92.9) | 80 (100) | 78 (95.1) |
| Grade 1 | 2 (2.4) | 0 (0) | 1 (1.2) |
| Grade 2 | 4 (4.7) | 0 (0) | 1 (1.2) |
| Grade 3 | 0 (0) | 0 (0) | 2 (2.4) |
| **Chills** | | | |
| No | 69 (81.2) | 70 (87.5) | 71 (86.6) |
| Grade 1 | 7 (8.2) | 9 (11.3) | 8 (9.8) |
| Grade 2 | 7 (8.2) | 1 (1.3) | 2 (2.4) |
| Grade 3 | 2 (2.4) | 0 (0) | 1 (1.2) |
| **Lymphadenopathy** | | | |
| No | 82 (96.5) | 77 (96.3) | 76 (92.7) |
| Grade 1 | 3 (3.5) | 2 (2.5) | 2 (2.4) |
| Grade 2 | 0 (0) | 0 (0) | 4 (4.9) |
| Grade 3 | 0 (0) | 1 (1.3) | 0 (0) |
| **Myalgia** | | | |
| No | 73 (85.9) | 61 (76.3) | 59 (72.0) |
| Grade 1 | 6 (7.1) | 11 (13.8) | 14 (17.1) |

| | SANOFI/GSK-D614 n=85 | SANOFI/GSK-B.1.351 n=80 | PFIZER BNT162b2 n=82 |
|---|---|---|---|
| | | n (%) | |
| Grade 2 | 6 (7.1) | 7 (8.8) | 8 (9.8) |
| Grade 3 | 0 (0) | 1 (1.3) | 1 (1.2) |
| **Nausea** | | | |
| No | 80 (94.1) | 76 (95.0) | 78 (95.1) |
| Grade 1 | 3 (3.5) | 3 (3.8) | 3 (3.7) |
| Grade 2 | 2 (2.4) | 1 (1.3) | 0 (0) |
| Grade 3 | 0 (0) | 0 (0) | 1 (1.2) |
| **Vomiting** | | | |
| No | 84 (98.8) | 80 (100) | 82 (100) |
| Grade 2 | 1 (1.2) | 0 (0) | 0 (0) |
| **Diarrhea** | | | |
| No | 81 (95.3) | 75 (93.8) | 75 (91.5) |
| Mild | 4 (4.7) | 4 (5.0) | 4 (4.9) |
| Moderate | 0 (0) | 1 (1.3) | 2 (2.4) |
| Severe | 0 (0) | 0 (0) | 1 (1.2) |
| **Pain in the extremities** | | | |
| No | 84 (98.8) | 77 (96.3) | 82 (100) |
| Mild | 1 (1.2) | 2 (2.5) | 0 (0) |
| Moderate | 0 (0) | 1 (1.3) | 0 (0) |
| **Insomnia** | | | |
| No | 82 (96.5) | 74 (92.5) | 79 (96.3) |
| Mild | 1 (1.2) | 5 (6.3) | 3 (3.7) |
| Moderate | 2 (2.4) | 0 (0) | 0 (0) |
| Severe | 0 (0) | 1 (1.3) | 0 (0) |

**Supplementary Figure 1.** Frequency of induction of various TH1 CD4$^{+}$ T-cell populations after a boost with BNT162b2 mRNA or Sanofi/GSK D614 or Sanofi/GSK B.1.351 vaccines.

After a boost with the various vaccines, a vaccine response for the different anti-SARS-CoV-2 TH1 CD4 subpopulations was considered significant if the ratio between D15 and D0 was >2 with a number of spots at D15 > 10 (after background subtraction).

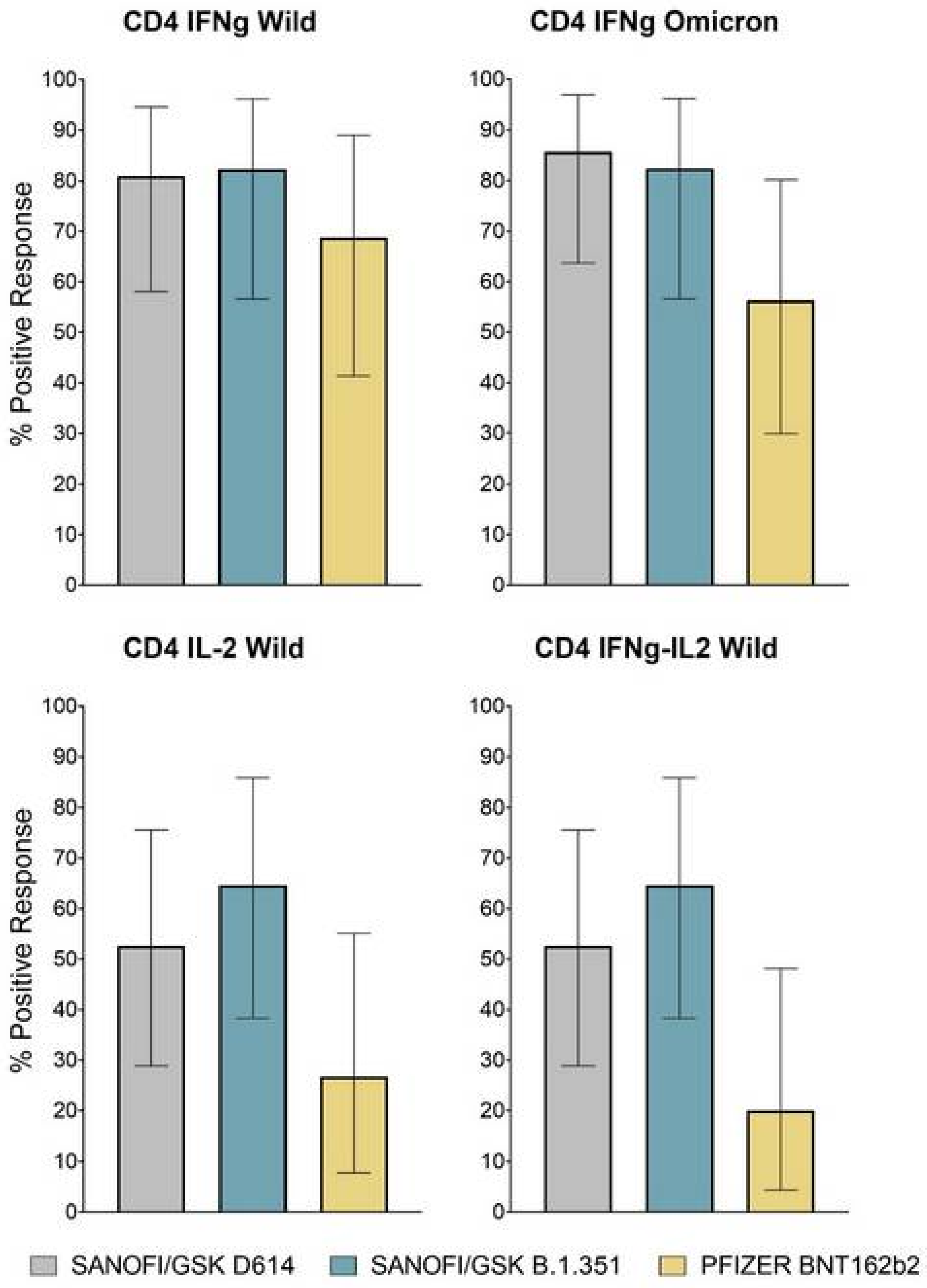
